# Optimizing Research Data Acquisition with Smart Pill Bottles (SPBs), the ORDAS Trial: A Feasibility and Implementation Study Protocol

**DOI:** 10.1101/2024.10.23.24315929

**Authors:** Nicolas Daccache, Joe Zako, Julien Burey, Olivier Verdonck, François Martin Carrier, Louis Morisson, Pascal Laferrière-Langlois

## Abstract

**Introduction:** Clinical trials are fundamental to advancing all areas of medicine. Despite their importance, trials are often expensive and time-consuming due to the need for extensive human resources, with limitations in cohort sizes and potential biases from loss of follow-up. Smart pill bottles (SPBs) offer a promising innovation by automating data collection, which could reduce costs and improve the granularity and accuracy of data. This technology may provide a more efficient alternative to traditional methods, streamlining data acquisition in clinical research.

**Objectives:** This proof-of-concept study aims to assess the feasibility of using smart pill bottles (SPBs) to collect data on opioid consumption in a postoperative setting, comparing their cost-efficiency and data quality to traditional methods. We hypothesize that SPBs will be readily adopted by users and enable the collection of highly granular data with fewer missing data points, while reducing the costs associated with human resource-based data collection.

**Material and Methods:** This single-center, single-arm trial will enroll 69 patients aged 18 and above undergoing major abdominal surgery via laparotomy. Following recruitment, patients will complete web-based questionnaires assessing pain, comorbidities, and quality of life. Postoperatively, patients will receive an SPB, the Thess Therapy Smart System manufactured by Thess Corporate (France) and provided by AppMed Inc. (Canada) to monitor opioid consumption at home for up to 90 days. At the end of the study period, participants will use the web-based platform to complete the same questionnaires, an opioid compliance checklist and a product satisfaction survey. The primary outcome will be the percentage of patients who successfully use the SPB throughout the study period. Secondary outcomes will include the extent of automated data collection, data granularity, project costs, the incidence of persistent opioid consumption, and patient satisfaction with the SPB.

**Trial registration:** clinicaltrials.gov (July 25th, 2024). Unique protocol ID: 2025-3801. NCT number: NCT06522698.

## 1. Background

Clinical trials are a fundamental component of clinical research, providing valuable evidence for assessing the efficacy and safety of medical interventions. In recent years, the number of published clinical trials has grown significantly, with estimates indicating that over 20,000 trials are initiated annually by drug developers. (1). This surge reflects the increasing reliance on rigorous scientific methods to inform medical practice and policy. While studies on large samples of patients are highly beneficial, a notable drawback lies in their time-consuming nature and the associated costs they incur. Moreover, the size of cohorts may be limited by the human resources and funds available, and loss of follow-up is an intrinsic risk to clinical trials that may introduce bias (2, 3). Despite the high costs, these investments are justified by the critical role that clinical trials play in advancing medical knowledge and improving patient outcomes. However, it is evident that developing tools to optimize the methods used in trials with the aim of increasing cost-efficiency would be highly beneficial given the substantial financial investments involved. Improved cost-efficiency could facilitate recruitment of larger patient cohorts and enhance the quality and diversity of research outputs assessed, ultimately leading to more robust and comprehensive findings. So how can we push the modernization of clinical research further, and what novel technologies could be used to streamline and automate data collection to increase cost-efficiency of research?

Several studies involving harnessing new technology to approach data collection have suggested that a streamlined and automated method of collecting data through connected technology can help set up clinical trials more cost-effectively (4, 5, 6). Evaluating the use of a connected device as a research tool in clinical trials and comparing it with traditional data collection using human resources would provide valuable insights into its efficiency and effectiveness. Smart medication adherence monitoring devices are a novel technology that provides objective and granular medication utilization data along with engaging patients with their treatment (7). Particularly, the smart pill bottle (SPB) is a rapidly developing technology that allows for medication monitoring of solid doses with the use of electronic sensors that can collect data on medication usage in real time and offer direct communication between patients and healthcare professionals or trialists (8). SPBs have shown efficacy in monitoring compliance and possibly increasing medication adherence in the clinical setting and the technology has been suggested as a potential research tool that would allow automatic collection of granular and precise data on the time of medication intake, dose, and frequency. (9, 10,). However, to our knowledge, there hasn’t been a trial comparing the efficacy of using SPBs for data collection in clinical trials versus the traditional method reliant on human resources in comparable contexts. Based on the properties of SPBs and available literature supporting the automatization and streamlining of data in clinical trials (4, 5, 6, 9, 10), we believe that the use of these devices may allow data collection of higher quality regarding granularity, number of losses of follow-up, completeness, missing data points along with a reduction of costs incurred by avoiding the use of human resources.

Earlier this year, the POCAS (Persistent opioid consumption after major abdominal surgery and its determinants) study was started at the University of Montreal. This prospective cohort study examined the prevalence of persistent opioid consumption (POC) three months post-surgery, its link to chronic postsurgical pain (CPSP), quality of life impacts, and risk factors. Researchers followed 668 patients, collecting opioid consumption data via telephone surveys and pharmacy reports, which required significant time and human resources. The POCAS trial is an example of a study in which the use of SPB could automate data acquisition, potentially reducing human cost while increasing the granularity of data (i.e. daily use vs monthly use).

This single group proof of concept study will mimic the POCAS trial methodology while incorporating SPB to explore its application in clinical research. More specifically, we aim to explore the level of adoption of SPBs in the context of postoperative research, describe the quality of data acquired, and describe the cost of data acquisition for a comparison with historical data.

## 2. Objectives and Hypotheses

Our primary objective is to evaluate the adoption of smart pill bottles (SPBs) in postoperative settings, to improve the process of data collection in the context of clinical trials.

Our secondary objectives will be to:

- Evaluate the quality of research data collected from patients regarding opioid consumption up to 90 days after major abdominal surgery with SPBs in the following metrics: automation of data collection, granularity of the data, number of missing data points.
- Evaluate the costs incurred for research data acquisition, using SPBs compared to standard practice using human resources, and compare these costs with historical cohorts.
- Estimate the prevalence of POC and CPSP 90 days after major abdominal surgery defined as the consumption of any quantity of opioids in the 7 days prior to the 90-day interrogation, describe its distribution in both opioid-naïve and chronic opioid consumers and across different types of abdominal surgery, and estimate its association with persistent pain and quality of life.
- Evaluate patient satisfaction with the use of SPBs.

Our exploratory objective is to explore how increased granularity of data collected in real time by the SPBs can enable deeper statistical analysis, in this specific case, build the trajectory of opioid consumption during the 90-day period to potentially identify patients at risk of developing chronic opioid consumption through predictive algorithms.

We hypothesized that:

- The implementation of SPBs as a data collection tool in clinical trials will be feasible, with patients expected to use the SPBs correctly at a rate of 80%, (11).
- The data generated from the SPB will be of high quality (increased granularity, lower missing data rate) as suggested by literature on similar products in clinical settings
- The costs involved in carrying out this project with the help of the SPBs will be lower than those incurred during the original POCAS study which collected data through human resources.
- The rate of POC will approximate 10%, as reported in the POCAS trial.
- Most patients will be satisfied with the use of the SPB, similarly to previously reported results (12).

## 3. Design and Methods

### a. Design

The project will be a one-arm single-site trial conducted at the CIUSSS-de-l’Est-de-l’Île-de-Montréal (Hôpital Maisonneuve-Rosemont).

### b. Population

The study population will consist of all consenting adult patients (age ≥ 18 years) undergoing major abdominal surgery via laparotomy (any open surgery involving the abdominal compartment or its wall, excluding appendectomies, inguinal hernia repairs, abdominal wall hernia repairs, and incisional hernia repairs). The population was chosen to resemble that of the POCAS study to ensure comparability.

Exclusion criteria include patients enrolled in the historical cohort (POCAS study) or currently participating in another study, patients undergoing additional surgery within 90 days post-hospital discharge, patient who will be oriented to a secondary care facility or rehabilitation centers and patients who do not understand French or English. We will also exclude patients with allergies or intolerance to hydromorphone, and for which hydromorphone is not the standard of care for postoperative pain management (see section below on Logistical Consideration).

### c. Outcomes

Our primary outcome will be the percentage of patients that will have appropriately adopted the SPB until the end of the 90-day period or until the absence of pain, defined as no hydromorphone intake for 72 hours. Non-adoption will be defined as the requirement to consume medication without the intended use of the SPB.

Our secondary outcomes will include:

- Automation of data collection, assessed by the number of human interventions required throughout the data collection process for each patient.
- Granularity of the data, assessed by the number of consumptions at each hour of every day.
- Total dose of opioid consumed (converted to daily oral morphine equivalent).
- Percentage of captured consumptions, assessed by the number of opioid capsules consumption properly tracked by the SPB, divided by the total number of capsules missing at the end of the protocol. In example, if the SPB tracked 18 capsules consumption, but 20 pills are missing from the container when retrieved, the percentage will be 90%.
- Percentage of questionnaires properly filled without interventions from the research staff.
- Percentage of patients who respected the initial prescription of opioid made by the physician upon hospital discharge. In example, if a patient received a prescription of hydromorphone 2 mg q4h PRN, and a delay below 4h between two consumptions was registered by the SPB, the patient will be deemed not to have followed the initial prescription.
- The costs incurred per patient from carrying out the project (research staff salaries, cost of device acquisition and daily use).
- Persistent opioid consumption (POC) 90 days after surgery as reported by the SPBs. POC will be defined as consumption of any quantity of opioids in the 7 days prior to the 90-day interrogation. This definition will be accurate for both preoperative chronic and non-chronic opioid users.
- The presence of CPSP in the 7 days prior to the interrogation. CPSP will be defined as any pain at the surgical site for patients who had pain at that site before surgery (by 1 point on the general numerical pain scoring question of the BPI questionnaire) (14).
- The change in the reported quality of life at the same time point. QOL will be measured as a continuous variable on the SF-12 questionnaire (15). These surveys have been selected to allow accurate comparison with the POCAS cohort as these were the surveys used in the context of that study.
- Patient satisfaction with the use of the SPB reported through a custom questionnaire

### d. Data Collection and Measurements

Once consent is obtained, we will complete the baseline interview with 5 questionnaires listed below in a period of approximately 20 minutes, either in person or by phone depending on availability. During this interview, we will gather baseline patient characteristics, including demographic information (age, sex, ethnicity, BMI, geographic location, economic status), medical and surgical history, preoperative medications, and the occurrence and location of preoperative pain. Chronic preoperative opioid usage will be determined as the intake of opioids for over three months before surgery (16).

We will use a modified short Brief Pain Inventory (BPI) questionnaire, adjusting it to inquire about pain over the previous 7 days instead of 24 hours, and replacing the body map question with a query about the pain site to distinguish surgical site pain from other pain locations. Additional questions in the questionnaire will address the chronic nature of preoperative pain. For chronic opioid users, we will collect data on the reasons for opioid use (if not solely for pain) and the duration of use.

We will assess comorbidities using the Charlson Comorbidity Index (CCI) (17). Baseline data on anxiety symptoms, depressive symptoms, catastrophizing tendencies, and quality of life will be collected using the PHQ-4, PCS-4, and SF-12 questionnaires, respectively (15, 18, 19). No intraoperative data will be recorded.

Following their surgery, all patients will be provided a SPB (Thess Therapy Smart System device, distributed by AppMed Inc, Québec, Canada), which will be used by patients to consume opioids and simultaneously record the consumption. Data regarding the dosage frequency, including the exact time of each consumption, will be continuously collected by the devices. During the 90-day post-op period, we will record any new hospitalizations, morbidity, or mortality.

If a patient’s medication consumption is not recorded by the SPB for 48 hours, they will automatically receive a notification inquiring about the reason for the cessation. More specifically, the patient will be asked whether the cessation is due to the absence of pain or another reason. If no answer is obtained within 24h, the research staff will directly contact the patient.

At the end of the 90-day post-op period, the patients will be prompted to use the proprietary web platform (Thess, France or p-Dose, AppMed Inc. Canada) to respond to end-of-trial surveys (PHQ-4, PCS-4, BPI, self-reported opioid consumption questionnaire, OCC checklist, SF-12 and patient satisfaction questionnaire). Once the SPB is retrieved, we will assess the number of capsules remaining in the container, and verify the integrity of the sticker, confirming that no medication was consumed outside of the normal dispensary mechanism.

### e. Patient recruitment and timeline

The research team will assess the elective surgical list of Maisonneuve-Rosemont Hospital’s operating room to identify eligible patients at least one week before their scheduled surgery.

Potential patients will be contacted by the research team via phone at least 3 days before surgery to provide an explanation of the project. The communication form used during these calls will be approved by the Maisonneuve-Rosemont Hospital Ethics Committee. Once consent is obtained and the consent form is adequately filled either via the REDCap application or via web signature (20), a thorough review of the patients’ medical charts will be conducted to confirm that they meet the inclusion criteria and have no exclusion criteria. Once eligibility is confirmed and patients are enrolled, the research team will meet the candidates before the surgery to complete the initial assessment, and to address any questions the patient may have. A first presentation of the SPB and how it works will be made by the research staff, but patients will be informed that a second demonstration will be completed prior to post-surgery hospital discharge.

After the surgery, once the patient recovered and there is imminent hospital discharge, the research staff will meet with the patient for a second demonstration of the SPB and the logistical consideration (SPB medication refill, retrieval at the end of the trial, etc.)

The 90-day monitoring period will start at the end of the final surgical procedure conducted during the initial hospital stay. This duration was selected because it aligns with the commonly accepted cutoff for identifying chronic persistent surgical pain, and is widely acknowledged in current research on postoperative patient outcomes and provides an advantageous window for potential preventive or preemptive interventions (21, 22, 23). It is also the time frame used in the POCAS study, facilitating the comparison with this historical cohort.

Throughout the 90-day monitoring period and depending on the consumption habit, the patient will be prompted to answer questionnaires on pain management via an automated system (i.e. email). If no answer is obtained within 24-48h, the research staff will directly contact the user to ensure questionnaire completion.

At the end of the 90-day monitoring period, the patient will be prompted to answer the final questionnaires and to return the SPB if it is still in his possession. If the patient still consumes opioids and needs an additional prescription, a consultation with his medical team will be organized to ensure the transition from the SPB use to standard prescription- and pharmacy-based medication dispense. Once these steps are completed, the patient’s research involvement will be completed. The timeline of events is illustrated in **Figure 1**.

**Figure 1.**
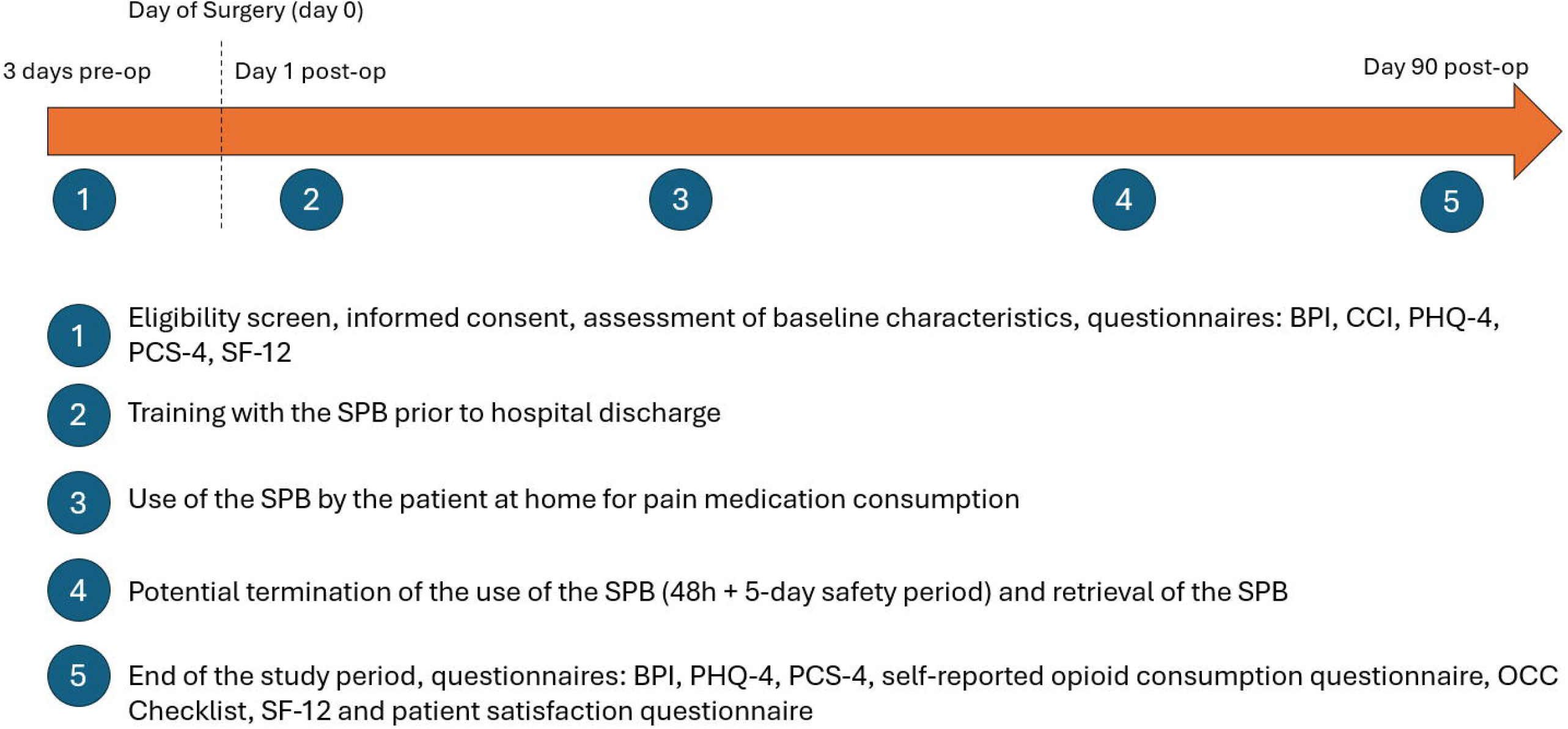
SPIRIT Schedule of Enrolment, Interventions and Assessments

We will recruit and collect data from patients at the CIUSSS-de-l’Est-de-l’Île-de-Montréal ((HMR) Hôpital Maisonneuve-Rosemont over a period of 12 months, starting in December 2024. A realistic estimate would be a recruitment of 3 patients per week over 23 weeks, which would amount to 69 patients included over approximately 5 months. A less favorable estimate would be a recruitment period of 9 months, followed by the 3-month period of follow-up and data collection. The rate of recruitment will be impacted by the number of SPB available, as provided by the commercial distributor. This work rate would allow the data analysis and submission of our initial results to be carried out within a 14-month frame.

### f. Description of SPB and logistical consideration

The SPB functions through a pressure sensitive cap that dispenses encapsulated medication when triggered. The SPB will provide medication without any dose and time restrictions, mimicking any standard container provided by a pharmacy. The patient will be instructed to solely request medication via the SPB dispenser mechanism and not to open the medication container. A sticker will be placed on the medication container to capture any instance of sticker tearing, indicating that the patient opened it.

Patients will also be instructed to present themselves back to the hospital center in the case of losing or breaking a smart pill bottle. A phone support line will be available to the patients 24 hours a day, 7 days per week for any problem with the hardware or software components.

Due to SPB current technological limitation, only encapsulated medication can be dispensed. Opioids are standardly not encapsulated, requiring us to manually encapsulate the medication use during this trial. The corollary of this limitation is that only the research pharmacy from Maisonneuve Rosemont hospital can provide the medication. Patients will initially be provided a dispenser with a maximum of 30 capsules and, and if needed, will have to contact the research staff and present themselves at the hospital to receive any refill.

Once patient has signaled no pain or medication requirement after being prompted by the research staff 48h following their last consumption, an additional period of 5 days without consumption will be given before retrieving the SPB. This additional period aims to account for any recurrence of pain and medication consumption, which would reinitiate the medication-free 48h required for completion of treatment. If no medication was requested for these 7 consecutive days, the research staff will retrieve the SPB, either via prepaid DHL packages sent to the hospital, or through the patient presenting themselves to the hospital.

### g. Data management

All data will be managed with respect to confidentiality and will not be used for any commercial purposes. A combination of software will be used throughout the patient timeline. For the initial consent of the patient, RedCap software application will be used. For further questionnaires, the commercial and proprietary WEB-based platforms will be used for data collection with the objective of simulating real-world applications of the SPB system. As further discussed below, the data collected by the commercial platform will be transferred to the research team for statistical analysis and data storage. It will not be used commercial development, and all copies held by the commercial partner will be erased upon termination of the research project.

To connect to the user interface web-based platform, the subject of the study will have to provide a valid e-mail and password. The later information collected will serve as identification to access the user interface WEB-based platform and for user id and password recovery. The login information will be stored on a separate secured database and serve only as login information control. The subject will be provided with a unique anonymized “*patient identification number*” by the clinical research coordinator. The research data collected by the user interface WEB-based platform and the medication dispensing device including the answers from the various questionnaires, the time and dosage of medication update and its corresponding adherence calculation will be associated to the “*patient identification number*”. No other information allowing to identify the subject will be collected and stored into the database (name, address, phone number, ect.). Only the clinical research coordinator will be knowledgeable of the association of the “*patient identification number*” to the subject *per se*.

The data will be automatically streamlined into a secured web-based electronic data collection system, without any intervention by the patient. The data collected will be anonymized and stored into dataset located on a Canadian-based highly secured server. At the end of the study, all anonymized data and database will be completely erased from AppMed’s systems and storage media. This destruction will be carried out securely to ensure that the data cannot be retrieved or used in the future. The user interface used to support the data collection into the database will consist of either *Thess* software (Thess, France) or *P-dose* software (AppMed inc., Quebec, Canada) depending on availability of the latter solution at the time of data collection. If the data recorded into the database is to be transferred from one database to another resulting from a change of user interface software, the procedure will be performed securely to mitigate any risks of data leaks and former database will be disposed according to the procedures described above. No private information or metadata, such as gelocalisation, will be extracted. All copies held by the commercial partner within their database will be erased upon termination of the research. No formal trial conduct auditing process will be performed.

Following the end of the trial, the research team will keep a copy of the data that will be retained for a period of 7 years. Paper copies (consent form) of patient information will be stored in a locked file cabinet in a locked office in the Department of Anesthesiology. Data will be assigned a unique study code which will be linked to the subject’s identities. The link will be kept separately from the data so that no patient can be identified in the event of loss or theft.

Every person involved in this study will receive appropriate training and abide by confidentiality guidelines to protect the subject’s privacy. All Health Information Protection Act rules and regulations will be strictly followed. The identification data will not be re-used or disclosed to a third party except as required by law, for authorized oversight of the research, or as permitted by an authorization signed by the research subject. The study has a low level of risk. The PI or designated personnel will regularly review all data, such as study completeness, enrollment, protocol deviations, dropouts, and adverse events, on a weekly or bi-weekly basis. Therefore, a data monitoring committee (DMC) will not be needed.

## 4. Data Analyses

### Sample Size

Based on available data in the literature concerning adherence to medication with the use of a SPB, we assume that we will estimate an adoption rate of 80%. This estimation is conservative when compared to Toscos et al. (11), who reported an adherence rate of 90% for patients using SPB for anticoagulant adherence. A precision of ± 10% will permit us to conclude that the observed adherence may be different than previously recorded ones. At a confidence level of 95%, 62 patients are required to have the precision needed on our adherence point estimate. To account for a potential dropout ratio of 10.0%, the number was inflated to 69 patients.

### Main Analyses

The descriptive analysis of the primary outcome and secondary categorial outcome will be presented as a frequency (%), which will be reported with the boundaries of the 95% confidence interval (CI). Continuous data will be presented using mean and standard deviation, or median and interquartile range if the distribution is skewed or not normal. According to the nature of the analyzed endpoints, 95% CI will be presented.

More specifically, we will conduct a descriptive analysis of the consumptions gathered per hour, per patient, the total opioid consumption for each patient (converted in daily oral morphine equivalents), the percentage of captured consumptions, assessed by the number of opioid capsules consumption properly tracked by the SPB, divided by the total number of capsules missing at the end of the protocol, the number of losses of follow up, the rate of response to the questionnaires on the WEB-based platform and the number of patients that needed to be reached by telephone, patient satisfaction with the product along with the costs incurred from carrying out the project. Missing data will be replaced by the maximum value observed (worst-case scenario).

When comparing with the historical cohort of POCAS, risk differences or mean differences with 95% confidence intervals will be reported for all outcomes, including the costs incurred by carrying out the project. The alpha value will be set at 0.05 to establish statistical significance. All data will be analyzed by the intention to treat principle. All statistical analysis will be performed using SAS, SPSS software or python/R programming language via VS Code or Jupyter notebook software.

We will also report metrics such as the SPB adoption rate and user satisfaction with the system stratified by patient demographics. For example, we will stratify results by age by splitting them at the median and comparing any differences. We will also conduct subgroup analyses on these outcomes by comparing opioid-naïve participants and opioid users along with the presence of any mental health condition reported in the initial comorbidity assessment. We will demonstrate the data collected on opioid consumption by the patients during the 90-day study period for each hour to provide an overview of the granularity of data.

For our exploratory objective, we will conduct a predictive analysis for the occurrence of POC, by using the trajectory of opioid consumption and the granular data of medication intake acquired throughout the 90-day period. We will explore machine learning and deep learning algorithms to predict POC at 90-days, while using the data collected as an input (demographics, time series of opioid consumption, etc.). The performance of the model will be assessed via *receiving operator characteristics* (ROC) curve and area under ROC (AUROC), precision, recall. Depending on the number of POC observed, we may amplify the true positive data by duplication. We will explore how newly crafted features (i.e. consumption of opioids at night) impacts the performance of the model. Finally, we will explore how the model performs at predicting POC, depending on the timing of prediction, throughout the 90-day period.

### Loss of follow-up

The number of patients lost to follow-up is expected to be limited due to the lending of the SPB, which will have to be retrieved. Patients who pass away before completion of treatment will be considered as lost to follow-up. In cases where they cannot be reached, it will be assumed that they have deviated from the proper use of the SPB, and they will be counted as non-adherent to the product. Subsequently, we will conduct several sensitivity analyses to investigate the implications of these lost-to-follow-up patients.

### Sensitivity Analyses

For our primary and secondary outcomes, we will use two models to explore the robustness of our findings and the impact of outcome measurement missingness due to the competing risk of death. We will fit models using the information gathered from proxies (spouse or family) or the medical chart of patients who have died by 3 months (potentially biased information) as well as one model with adherence amputated on those patients and one amputating no adherence on the same patients (best- and worst-case scenarios analyses).

## 5. Ethical Considerations

The study will adhere to the protocol, the Declaration of Helsinki, Good Clinical Practice (GCP), and relevant Canadian laws and regulations. This protocol has been approved by CER- CEMTL (CIUSSS de l’Est-de-l’Île-de-Montréal) Research Ethics Board (REB). Any amendments to the project will also undergo REB approval. Furthermore, the Thess Therapy Smart System will have undergone approval for patient use by Health Canada before the start of the trial.

Prior to starting the study, patients will be contacted by telephone at least 3 days before their surgery. We will describe the project and, if participation interest is confirmed, ask them to consent to data collection preoperatively and postoperatively through the Thess device for a 3- month period. In cases where patients are unable to provide an electronic signature via RedCap, we will obtain verbal consent during our first contact by telephone and finalize the consent process by obtaining written consent the day of the surgery. For patients we meet on the day of surgery, we will secure written consent before proceeding with preoperative interviews, whether conducted by phone or in person.

We will request a consent model from CIUSSS de l’Est-de-l’Ile-de-Montreal’s REB that incorporates Article 3.7A of the TCPS2 concerning modifications to the consent process and partial disclosure. Patients will be made familiar with the Thess Therapy Smart System device and will be informed of the primary objective of this study which is to evaluate the capabilities of the device in data collection. They will be instructed to use the device only for medication consumption to avoid erroneous inputs. We will inform patients that the study focuses on the use of SPBs for medication intake without disclosing the secondary objective, which is measuring persistent opioid consumption. We believe that disclosing the objective could lead to a conscious change in behavior related to medication consumption and introduce bias. This partial disclosure may improve data accuracy regarding patients’ opioid use, and secondary comparison with the historical cohort of POCAS trial. We will ensure that patients do not feel inhibited or judged about their potential opioid consumption, as it will be presented as a potentially normal behavior three months after surgery. Following completion of the study, after having completed all questionnaires, participating patients will be contacted by phone call to explain the secondary purpose of the study and the rationale behind the initial partial disclosure as described above. Following this summary, patients will be asked if they still consent to participate in the study knowing its full scope and if not, the data will be destroyed. We believe that fully disclosing the secondary objective upfront would impede our ability to accurately measure the secondary outcome.

Patient information collected either through chart review, through surveys on the web platform or automatically through the SPB device will be securely stored for a period of 7 years on a protected server within the research lab of Maisonneuve-Rosemont hospital, maintaining strict confidentiality. This data will not be used in ancillary studies. Each patient will receive a unique trial identification number to safeguard anonymity, which will correspond to allocation data in secure electronic records. Research personnel will be instructed to segregate any patient data containing personal identifiers, storing them under their respective ID numbers. The trial master file will be securely maintained at the study site throughout the research period. No data allowing the identification of study participants will be revealed in any publications, ensuring their anonymity. Any individual medical data obtained during the study will be treated as confidential and not disclosed to third parties except under specific circumstances:

- Medical information might be shared with the subject’s personal physician or other relevant medical professionals overseeing the subject’s care with the patient’s consent.
- The data resulting from the study will be accessible for inspection upon request by participating physicians, Research Ethics Boards (REBs), and regulatory authorities.

Patients experiencing significant pain will have the option for a telephone follow-up aimed at assisting them in accessing suitable care. If the principal investigator observes that pain remains uncontrolled despite necessary follow-up, the patient will be offered a consultation at the HMR Pain Clinic. Since patients will be queried about opioid-related issues, our aim is to motivate them to seek assistance when necessary and to provide referrals to appropriate resources in cases of psychological distress associated with opioid use. Furthermore, interventions may be warranted in cases where significantly high intake of opioid is reported by a patient’s Thess device.

## 6. Feasibility

### Financial aspects

The SPBs will be provided by the distributor at no cost. However, as the SPBs only function using encapsulated medication for the moment, expenses associated with encapsulating opioid pills must be considered. As most patients typically require opioid medication for only a few days post-surgery (24), the quantity of pills requiring encapsulation will remain economically feasible. Despite the level of automation involved in the data collection, the most important expenses will be linked to the human resources required. These expenses which will be paid from active grants accounts.

## 7. Strengths and Limitations

This study will help evaluate smart pill bottles as data collection tools through an accurate comparison in time and context with the classical method reliant on human resources. Positive findings may introduce a revolutionary data acquisition method that would open the door to large-scale projects involving large cohorts of patients, while benefiting from a reduction in associated costs. A significant amount of resources is dedicated to clinical research, but few studies seek to improve the research processes themselves, and to objectively document the improvement of these processes. In this sense, this innovative project has the potential to modernize research data collection strategies, demonstrating an advantage in terms of both quality and cost.

Our study may be limited due to the nature of the technology that records medication intake through sensors that monitor the number of times the pill box was opened rather than the true amount of consumed medication. This design characteristic may lead to inaccuracies in the data reported by the devices. Additionally, the device’s dependence on encapsulated medication could result in escalated expenses due to the encapsulation procedure. While this cost is specific to the study’s focus on opioids and the device’s operational constraints, we will need to discuss the potential cost advantages that would arise from addressing this challenge. Furthermore, the nature of the treatment regimen of our cohort (medication to take as needed) may make an adherence analysis more challenging than with a scheduled treatment, as patients may stop taking the medication without indicating the reason through the questionnaires.

Moreover, the study’s duration and the restricted frequency of patient assessments in the POCAS study might underestimate the potential cost savings generated by implementing Smart Pill Bottles (SPBs). In research requiring multiple contacts with participants, the cost differential between human resource-based data collection and SPB automated monitoring could be substantially more pronounced.

## Data Availability

No datasets were generated or analyzed during the current study. All relevant data from this study will be made available upon study completion.
Any additional data such as the patient consent form or questionnaires will be made available upon reasonable request to the corresponding author.

## 9. Access to data

Only the PI, the co-investigators and the research staff will have access to all the data during the study and study analysis.

## 10. Dissemination policy

We plan to publish the results of this trial in a high-impact factor journal in the field of anesthesia, or medical technology.

Patients or the public were not involved in the design, or conduct, or reporting, or dissemination plans of our research. Full protocol as well as supplementary data from this study could be made available upon request to the PI. To be eligible for authorship, contributors must have significantly participated in the creation, development, or enrollment of participants or statistical analysis of the study.

We plan on granting public access to the full protocol through publication. Participant-level dataset and statistical code can be made available upon reasonable request to the corresponding author.

## 11. Author Contributions

Conceptualization – ND, JB, JZ, LM and PLL; Investigation – ND, JB and PLL; Methodology – ND, JB, JZ, LM, FMC, OV and PLL; Supervision – PLL; Writing (Original Draft Preparation) – ND and JB; Writing (Review & Editing) – ND, LM and PLL.

## 12. Acknowledgements

Thanks to all the research team at LIAM (Laboratory of innovative anesthesia in Montreal) for its help in the research organization.

## Appendix 1 Trial Registration Dataset

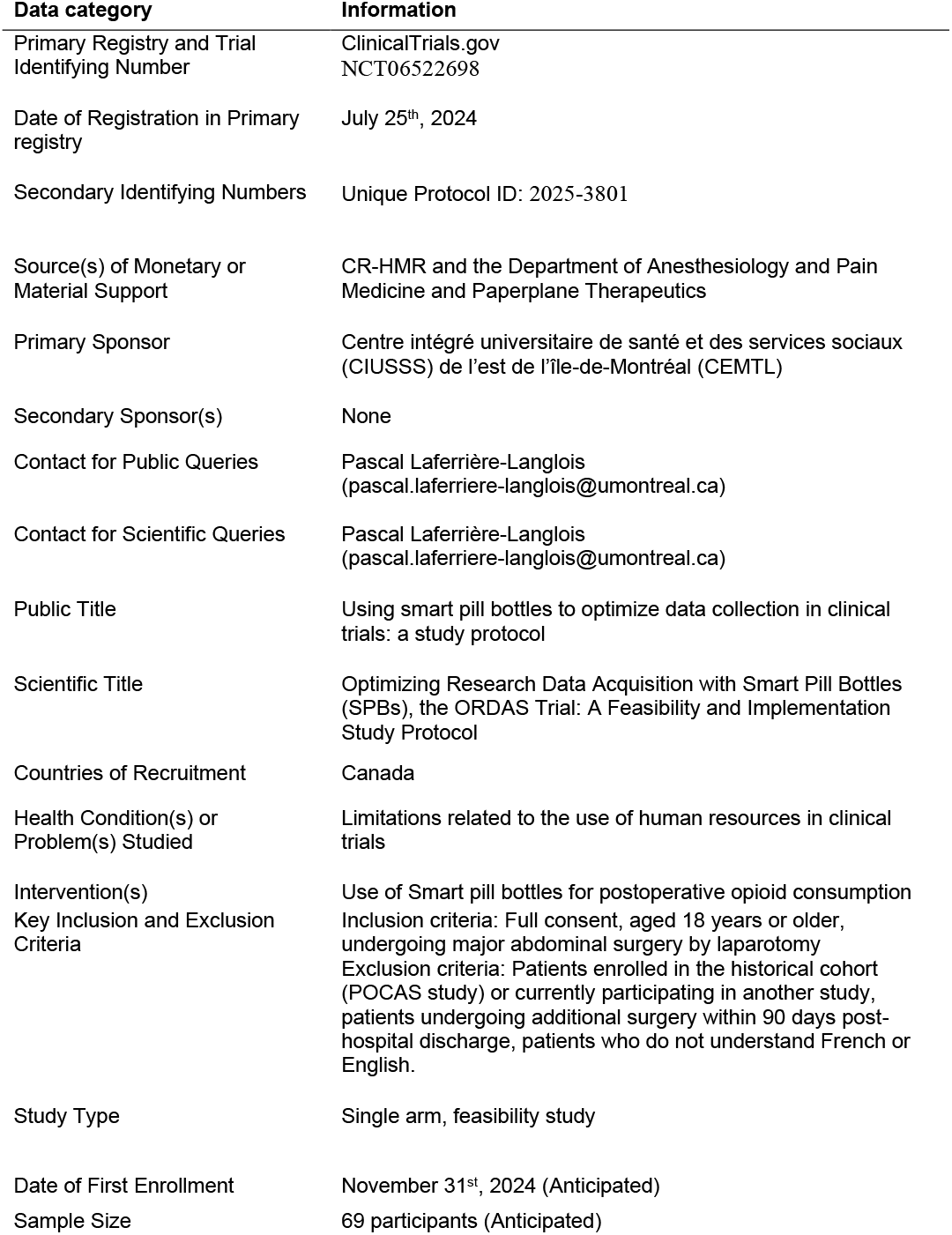
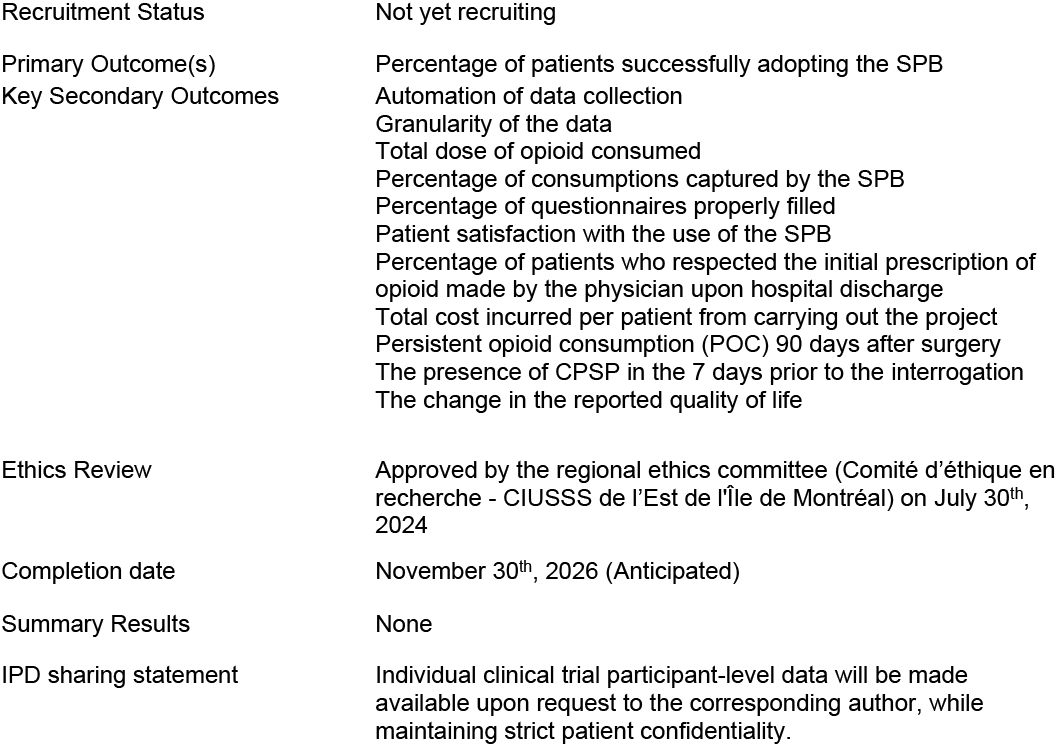

